# Prevalence Of carbapenem resistance in *Acinetobacter baumanii* and *Pseudomonas aeruginosa* in sub-Saharan Africa: a systematic review and meta-analysis

**DOI:** 10.1101/2022.11.29.22282516

**Authors:** Margaret Toluwalayo Arowolo, Oluwatosin Qawiyy Orababa, Morufat Oluwatosin Olaitan, Bisola Victoria Osibeluwo, Utibeima Udo Essiet, Olajumoke Hanah Batholomew, Oluwaseyi Gbotoluwa Ogunrinde, Oluwaseyi Aminat Lagoke, Jeffrey Difiye Soriwei, Olanrewaju David Ishola, Onyinye Maryann Ezeani, Aminat Oyeronke Onishile, Elizabeth Olumodeji

**Affiliations:** Department of Microbiology, University of Lagos, Akoka, Lagos, Nigeria; School of Life Sciences, University of Warwick, Gibbet Hill Campus, Coventry, United Kingdom; Department of Microbiology, University of Ibadan, Ibadan, Nigeria; School of Health Sciences, University of Bristol, United Kingdom; Department of Public Health in Microbiology, University of Bedfordshire, Luton, United Kingdom; Department of Biomedical Science, Nottingham Trent University, United Kingdom; Faculty of Health Studies, School of Nursing and Healthcare Leadership, University of Bradford, West Yorkshire, England, United Kingdom; Department of Biology, Texas Women University, United States

**Keywords:** Antibiotic resistance, *Acinetobacter baumannii*, *Pseudomonas aeruginosa*, sub-Saharan Africa, carbapenem-resistant *A. baumannii*, carbapenem-resistant *P. aeruginosa*

## Abstract

**Background:** Carbapenems are drugs of last resort and resistance to them is considered a great public health threat, especially in notorious nosocomial pathogens like *Acinetobacter baumannii* and *Pseudomonas aeruginosa*. In this study, we aimed to determine the prevalence of carbapenem resistance in *A. baumannii* and *P. aeruginosa* infections in Sub-Saharan Africa.

**Methods:** Databases (PubMed, Scopus, Web of Science, and African Journal Online) were systematically searched following the Preferred Reporting Items for Systematic review and meta-analysis protocols (PRISMA-P) 2020 statements for articles reporting carbapenem-resistant *Acinetobacter baumannii* (CRAB) and carbapenem-resistant *Pseudomonas aeruginosa* (CRPA) prevalence between 2012 and 2022. Pooled prevalence was determined with the random effect model in R.

**Results:** A total of 47 articles were scanned for eligibility, among which 25 (14 for carbapenem-resistant *A. baumanii* and 11 for carbapenem-resistant *P. aeruginosa*) were included in the study after fulfilling the eligibility criteria. The pooled prevalence of CRPA in the present study was estimated at 8% (95% CI; 0.02 – 0.17; I^2^=98%; P <0.01). There was high heterogeneity (Q=591.71, I^2^=98.9%; P<0.0001). The pooled prevalence of CRAB in the present study was estimated at 20% (95% CI; 0.04 – 0.43; I^2^=99%; P <0.01). There was high heterogeneity (Q=1452.57, I^2^=99%; P<0.0001). Carbapenem-resistant *A. baumannii* prevalence based on sample source gave estimates of 24% (95% CI; 6 – 49; I^2^=99%; P<0.01). The carbapenamse genes commonly isolated from *A. baumanii* in this study include *bla*_OXA23,_ *bla*_OXA48_, *bla*_GES._, *bla*_NDM,_ *bla*_VIM_,, *bla*_OXA24_, *bla*_OXA58_, *bla*_OXA51_, *bla*_SIM-1_, *bla*_OXA40_, *bla*_OXA66_, *bla*_OXA69_, *bla*_OXA91_, with *bla*_OXA23_ and *bla*_VIM_ being the most common. On the other hand, bla_NDM,_ bla_VIM_, bla_IMP_,, bla_OXA48_, bla_OXA51_, *bla*_SIM-1_, *bla*_OXA181_, *bla*_KPC_, *bla*_OXA23_, *bla*_OXA50_ were the commonly isolated carbapenemase genes in *P. aeruginosa*, among which bla_VIM_ and bla_NDM_ genes were the most frequently isolated.

**Conclusion:** Surveillance of drug-resistant pathogens in sub-Saharan Africa is essential in reducing the disease burden in the region and this study has shown that the region has significantly high multi-drug resistant pathogen prevalence. This is a wake-up call for policymakers to put in place measures to reduce the spread of these critical priority pathogens.

## INTRODUCTION

Antimicrobial resistance (AMR) is a leading public health threat globally, considerably escalating morbidity, mortality and treatment failure of microbial infections, as well as economic losses to individuals and nations (Prestinaci et al., 2015). According to the 2016 Review on Antimicrobial Resistance, AMR would be responsible for 10 million deaths yearly by 2050 with a large amount of these deaths occurring in Sub-Saharan Africa (O’Neil, 2016). However, a recent report of almost 5 million deaths linked with AMR in 2019 alone has shown that we will be reaching the 10 million AMR-associated deaths sooner than earlier predicted (Murray et al., 2022).

Carbapenems are beta-lactam antibiotics with broad-spectrum bactericidal activities against both Gram-positive and negative pathogens (Armstrong et al., 2021). They are often used as last-line drugs against bacterial infections (Ma et al., 2021). Unfortunately, bacterial pathogens have developed resistance to this last-resort group of antibiotics through various genetic modifications and production of carbapenem-hydrolyzing enzymes (Meletis, 2016). This has made infection therapy more difficult and expensive, especially against the notorious Gram-negative bacteria.

*Acinetobacter baumanii* and *Pseudomonas aeruginosa* are Gram-negative pathogens that belong to the ESK**AP**E group. These are bacterial pathogens that are notorious for their resistance to clinically relevant antimicrobials. Furthermore, carbapenem-resistant *A. baumanii* and *P. aeruginosa* have both been grouped as critical priority pathogens for which there is a need to develop new and effective antimicrobials. These have made the surveillance of these two pathogens a necessity, especially in low-middle income countries where surveillance is poor.

Sub-Saharan Africa is known for having a high burden of infectious diseases which might be linked to the poverty level and poor water, sanitation, and hygiene (WASH) practices in the region (Whiteside and Zebryk, 2017; Zerbo et al., 2021). The Murray et al. (2022) report also showed that sub-Saharan Africa suffers the highest prevalence of AMR-associated death globally. The scarcity of AMR data from SSA has made it difficult to determine the true risk and burden of AMR infections in the region. Also, a recent study predicting the global prevalence of carbapenemase-producing *P. aeruginosa* could not include majority of sub-Saharan African countries in the analysis due to scarcity of data. This further emphasizes the need for surveillance of AMR pathogens in Africa, most especially sub-Saharan Africa. To the best of our knowledge, there is currently no up-to-date systematic review and meta-analysis that reports the pooled prevalence of *P. aeruginosa* and *A. baumanii* in sub-Saharan Africa. This systematic review is an extensive analysis of the prevalence of carbapenem-resistant *Acinetobacter baumannii* with the aim to describe the epidemiology within sub-Saharan Africa. This study also investigated the prevalence of carbapenemase genes in the region. This would provide insight on the public health risks posed by these priority pathogens and the development of sustainable prevention and control interventions in this region.

## METHOD

### Search Strategy

This study was performed according to the Preferred Reporting Items for Systematic Review and Meta-analysis (PRISMA) 2020 guidelines (Page et al., 2021). Electronic databases (PubMed, Scopus, Web of Science, and African Journal Online) were searched for articles published on carbapenem-resistant *A. baumanii* and/or *P. aeruginosa* in sub-Saharan Africa. The last search was on the 31st of July 2022.

### Eligibility Criteria

This study only included studies published in sub-Saharan Africa between 2012 and 2022. Only studies that reported *A. baumanii* and/or *P. aeruginosa* prevalence in humans were considered for the analysis. Studies that reported *A. baumanii* and/or *P. aeruginosa* prevalence from countries outside sub-Saharan Africa were excluded from the analysis. Also, only studies that reported at least one case of carbapenem resistance in *A. baumanii* and/or *P. aeruginosa* were included. Lastly, cross-sectional studies, retrospective, and longitudinal analyses were considered in this analysis. Literature and systematic reviews were excluded from this analysis.

### Screening Strategy

Articles were first screened based on their titles and abstracts by two independent researchers. Two other researchers then screened eligible articles by reading through their full text, ineligible articles were excluded and the reasons for their exclusion were stated (Figure 1). All disagreements were resolved through discussions. A third researcher confirmed the eligibility of the included studies before including them in the analysis. During extraction, for studies that reported different percentage for imipenem or meropenem, preference was given to meropenem since it has better activity against Gram-negative bacteria (Zhanel et al., 1998).

**Figure 1.**
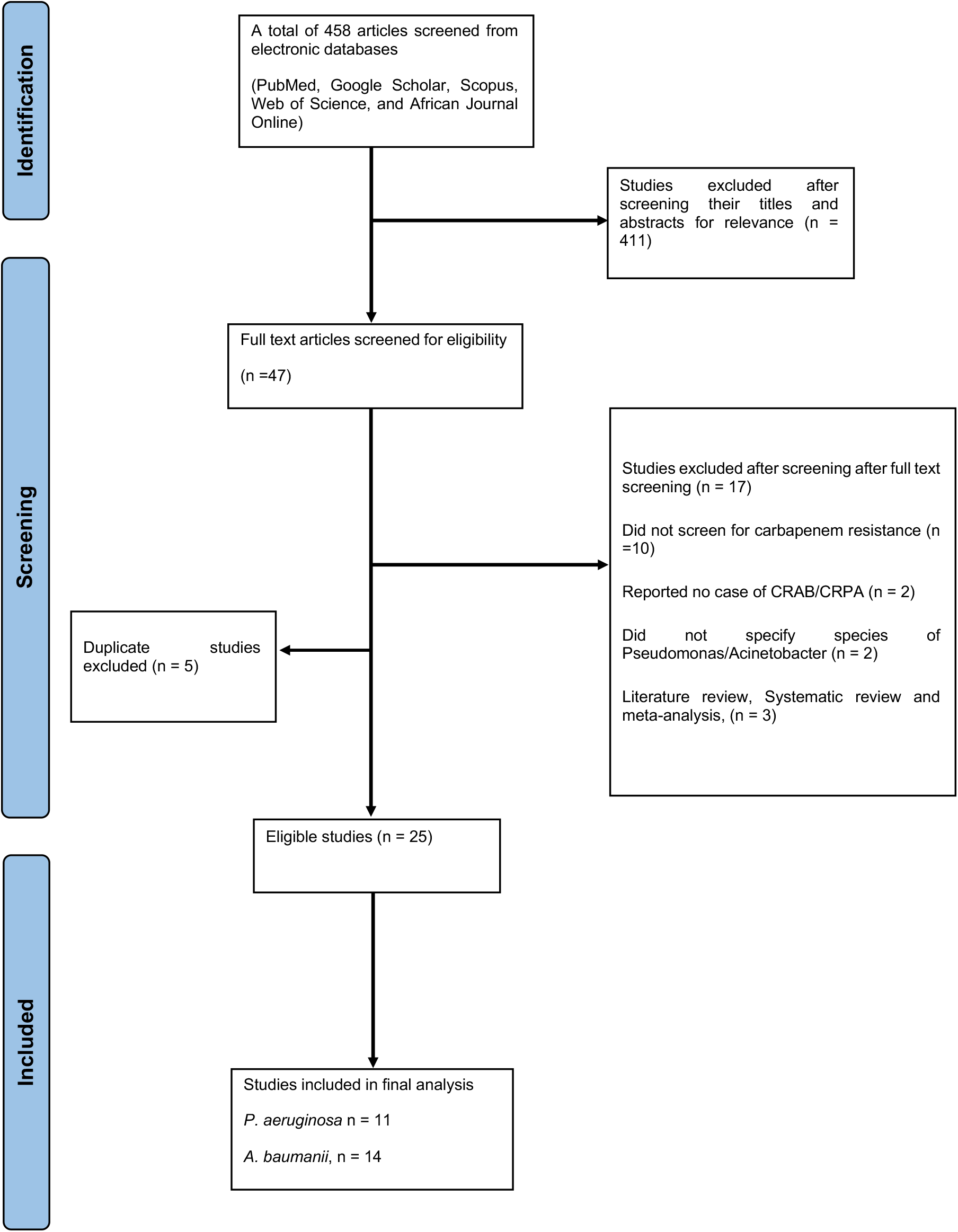
PRISMA flowchart for the search process.

### Data Extraction and Quality Appraisal

A data extraction table was created by one reviewer and two other reviewers extracted the following data from the eligible articles; first author, study design, study country, study aim, sample size, sample source, organism prevalence, carbapenem-resistance prevalence, AST method, carbapenem used. All disagreements were resolved through discussions and a fourth reviewer confirmed the extracted data. This step was performed for each of the organisms.

### Data Analysis

The data extracted was cleaned for any eligibility criterion error, and the meta-analysis was performed using RStudio version 4.2.0. The Carbapenem-resistant *A. baumannii* (CRAB) and *P. aeruginosa* (CRPA) pooled prevalence of the present study was analyzed using the random-effect model while subgroup prevalence was considered based on the source of the reported studies. Cochran Q statistics and the I^2^ (inverse variance index) were used to analyze the heterogeneity (low, 0-0.25; fair, 0.25-0.5; moderate, 0.5-0.75; and high, above 0.75). A p-value of <0.01 was considered significant. Publication bias was examined visually with the use of the funnel plot.

## RESULT

A total of 25 articles (CRAB, 14; CRPA 11) (Figure 2) were included in the final analysis after screening 458 articles from different electronic databases. These articles were from different countries and sub-regions of sub-Saharan Africa. Of the 14 articles analysed for the CRAB, 4(28.6%) were from South Africa, Sudan and Uganda had 2(14.3%) each, while Ethiopia, Senegal Malawi, Kenya, Nigeria, and Sierra Leone had one each. For the CRPA analysis, Nigeria, Ethiopia, and Sudan each had 2 eligible articles included in the final analysis while Malawi, Ghana, and Uganda each had just one eligible article.

**Table 1.**
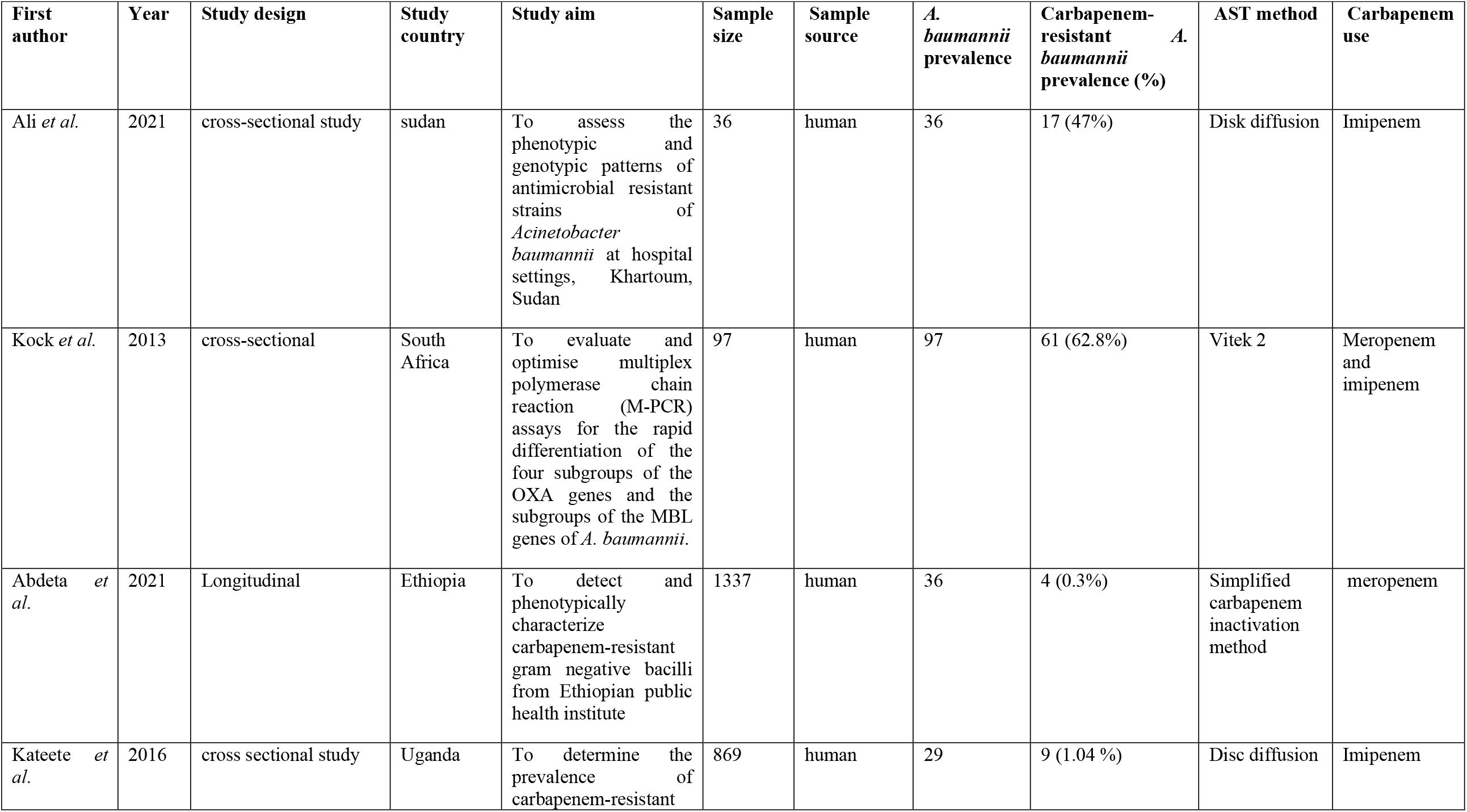
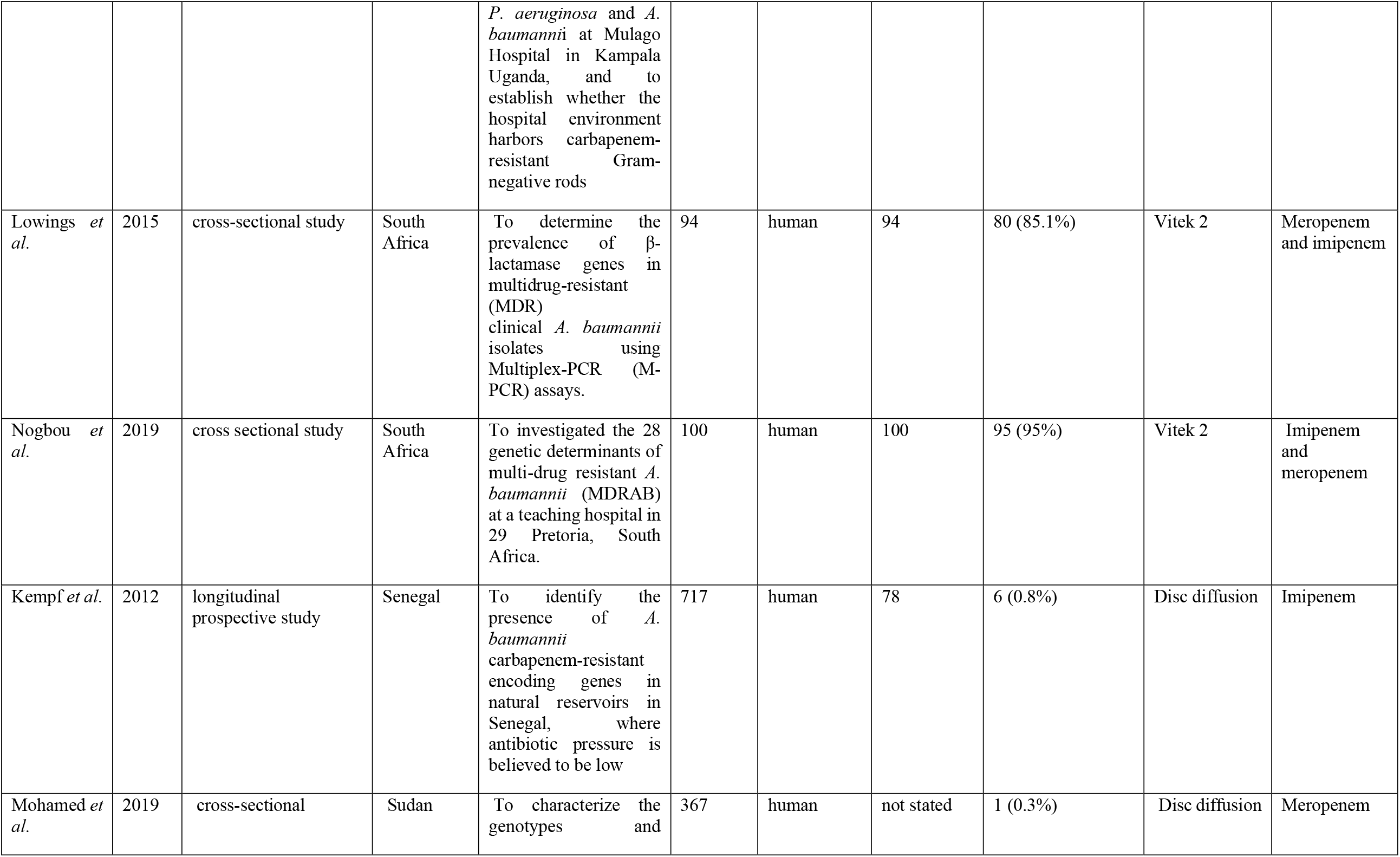
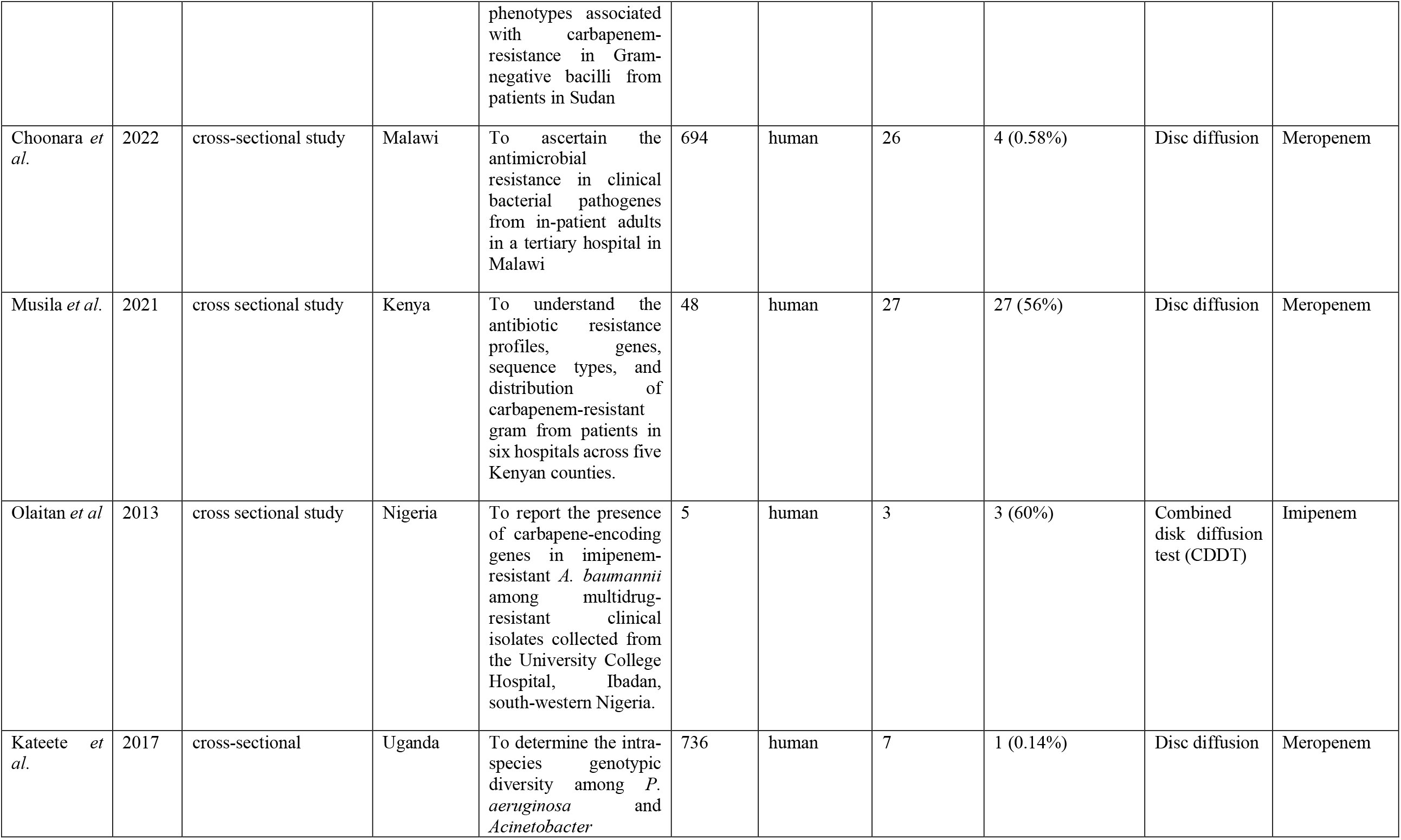
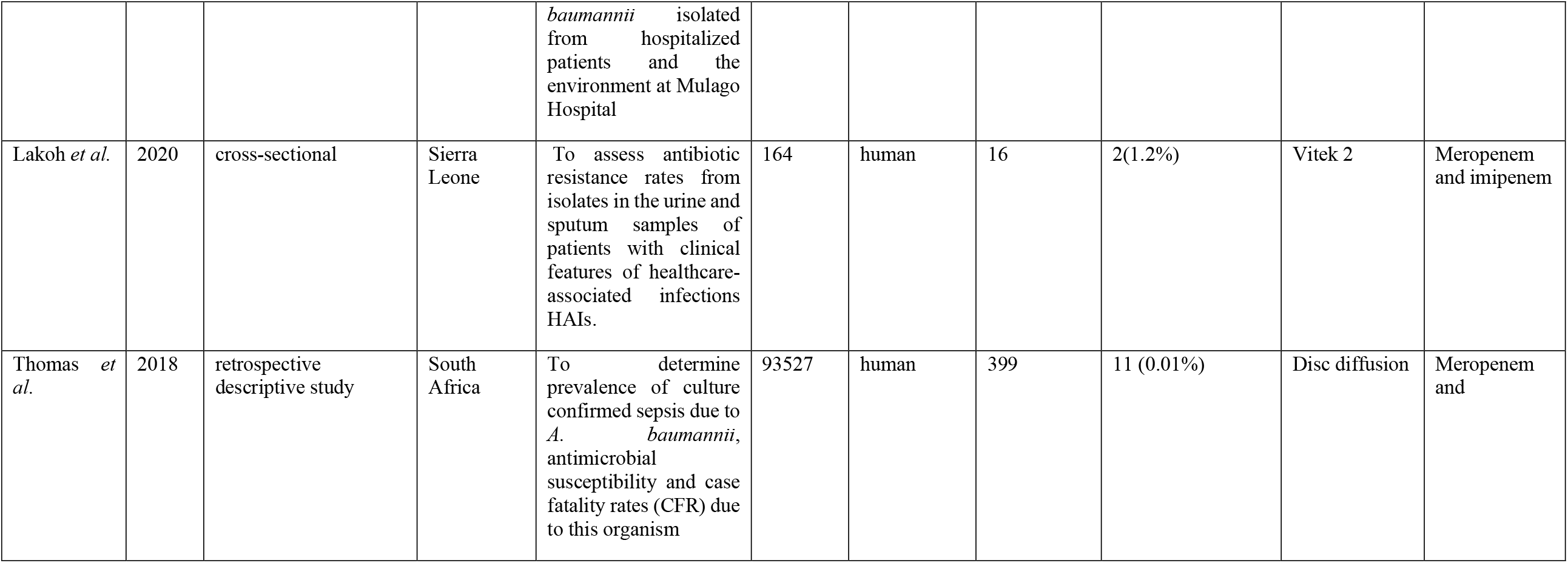
Data extraction table for carbapenem-resistant *A. baumannii* studies.

**Figure 2.**
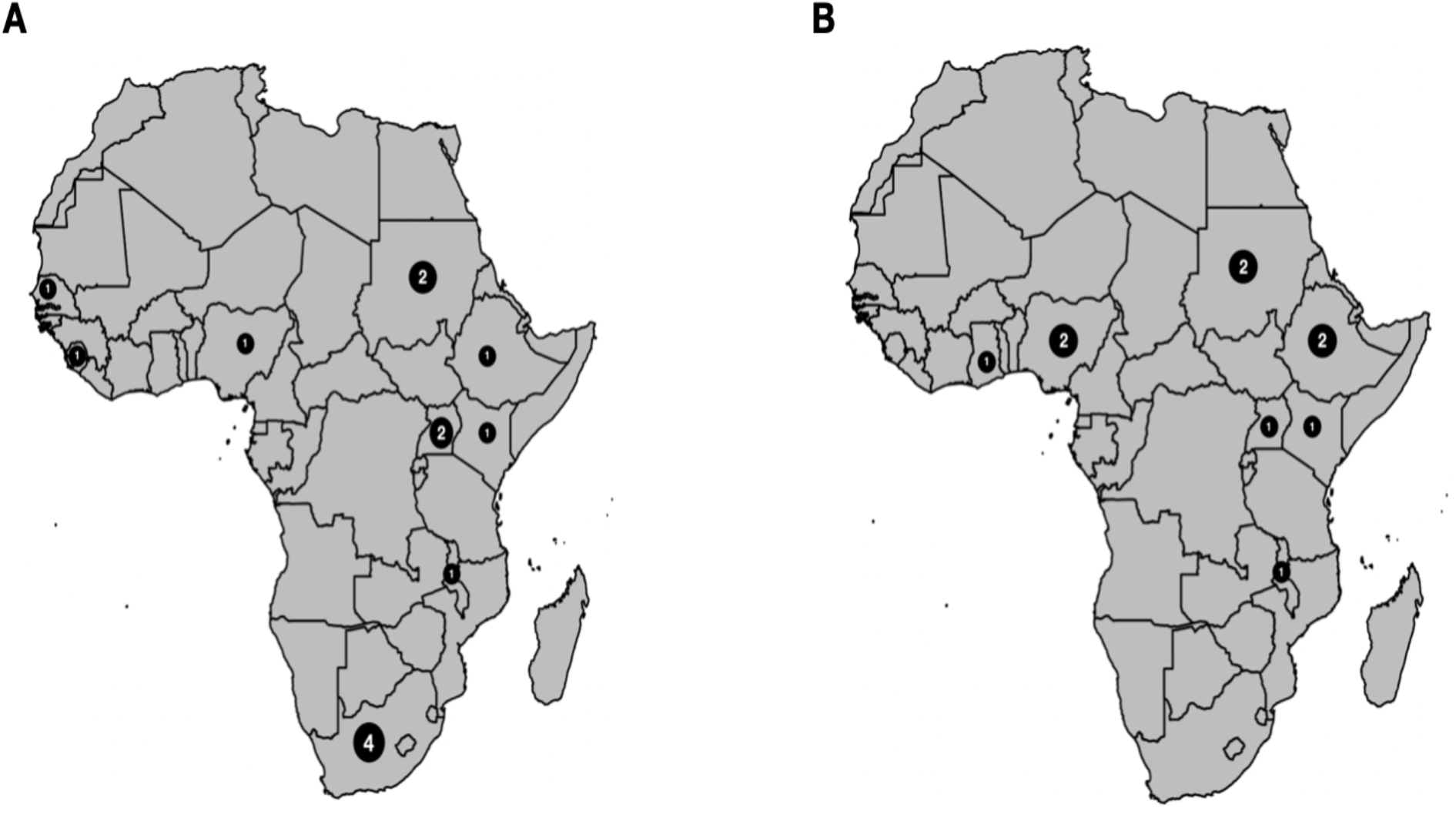
Distribution of eligible articles included in the final analysis. A. A total of 14 articles were analysed to determine the pooled prevalence of carbapenemen-resistant *Acinetobacter baumanii*. Majority (4) were from South Africa, Sudan and Uganda had 2 each, while Ethiopia, Senegal, Malawi, Kenya, Nigeria, and Sierra Leone had one each. B. A total of 11 articles were analysed to determine the pooled prevalence of carbapenemen-resistant *Pseudomonas aeruginosa*, 2 each from Ethiopia, Nigeria, and Sudan while Malawi, Ghan, Kenya, and Uganda had one each.

### Meta-analysis of Carbapenem-resistant *A. baumannii*

The pooled prevalence of CRAB in the present study was estimated at 20% (95% Confidence Interval [CI]; 0.04 – 0.43; I^2^=99%; P <0.01) (Figure 3). The Q statistics shows high heterogeneity (Q=1452.57, I^2^=99%; P<0.0001) between the CRAB studies.

**Figure 3.**
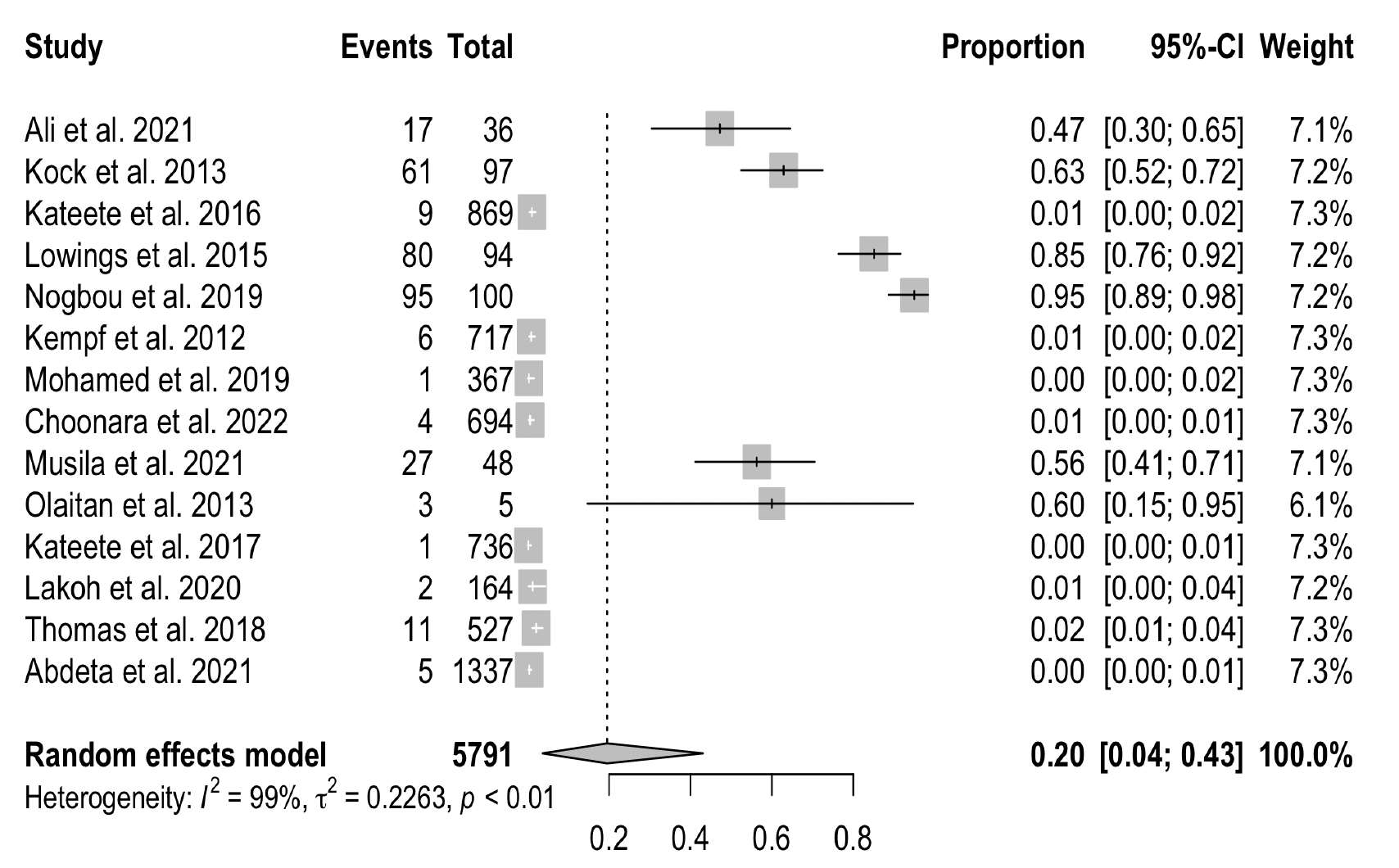
The Forest plots of random-effects meta-analysis show the pooled prevalence of carbapenem-resistant *Acinetobacter baumannii*. CI= Confidence interval

### Meta-analysis of Carbapenem-resistant *P. aeruginosa*

The pooled prevalence of CRPA in the present study was estimated at 8% (95% CI; 0.02 – 0.17; I^2^=98%; P <0.01) (Figure 4). Similarly, there was high heterogeneity (Q=591.71, I^2^=98.9%; P<0.0001) between CRPA studies analysed.

**Figure 4.**
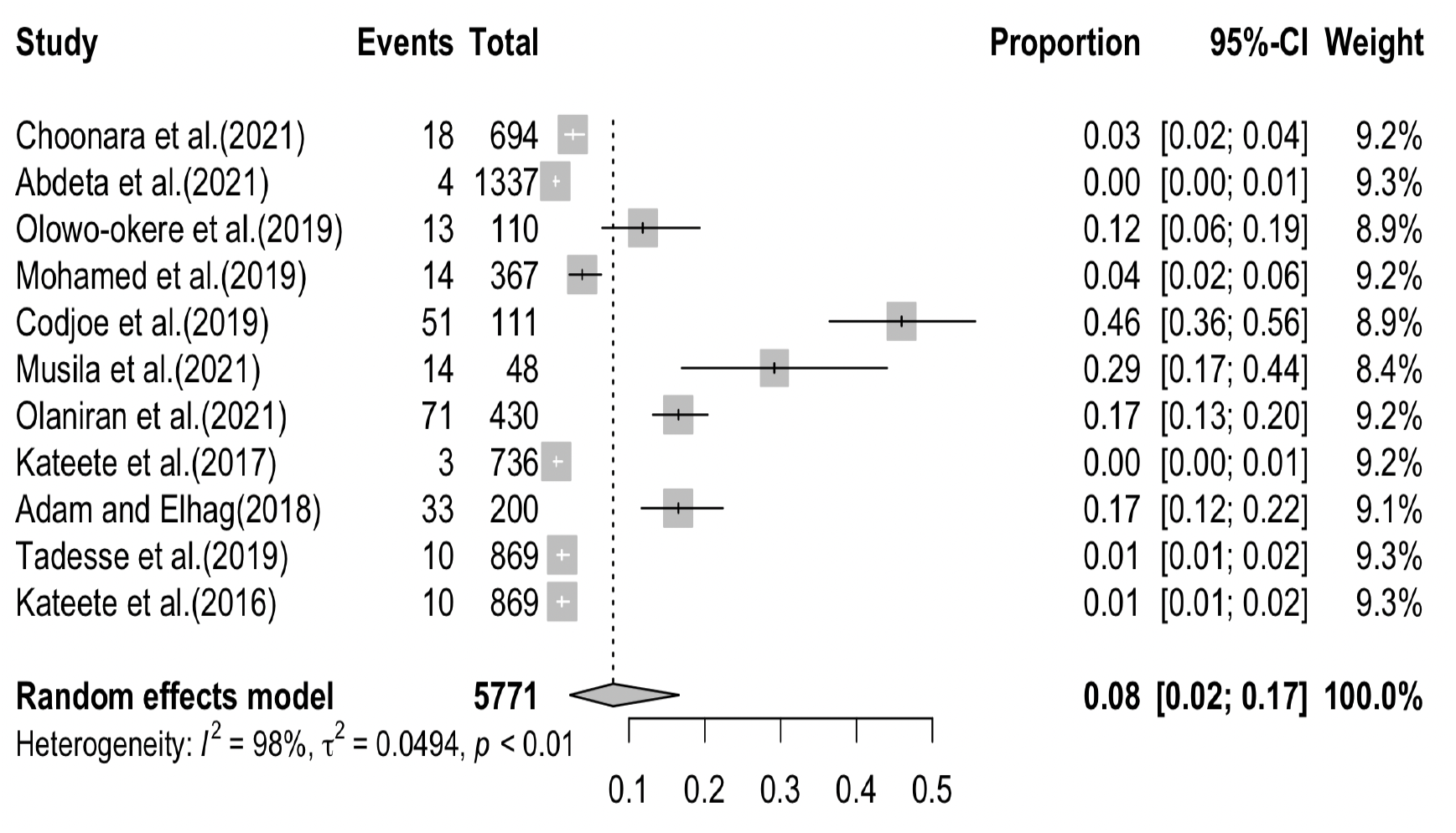
Forest plot showing the pooled prevalence of carbapenem-resistant *Pseudomonas aeruginosa* in sub-Saharan Africa.

**Fig. 5.**
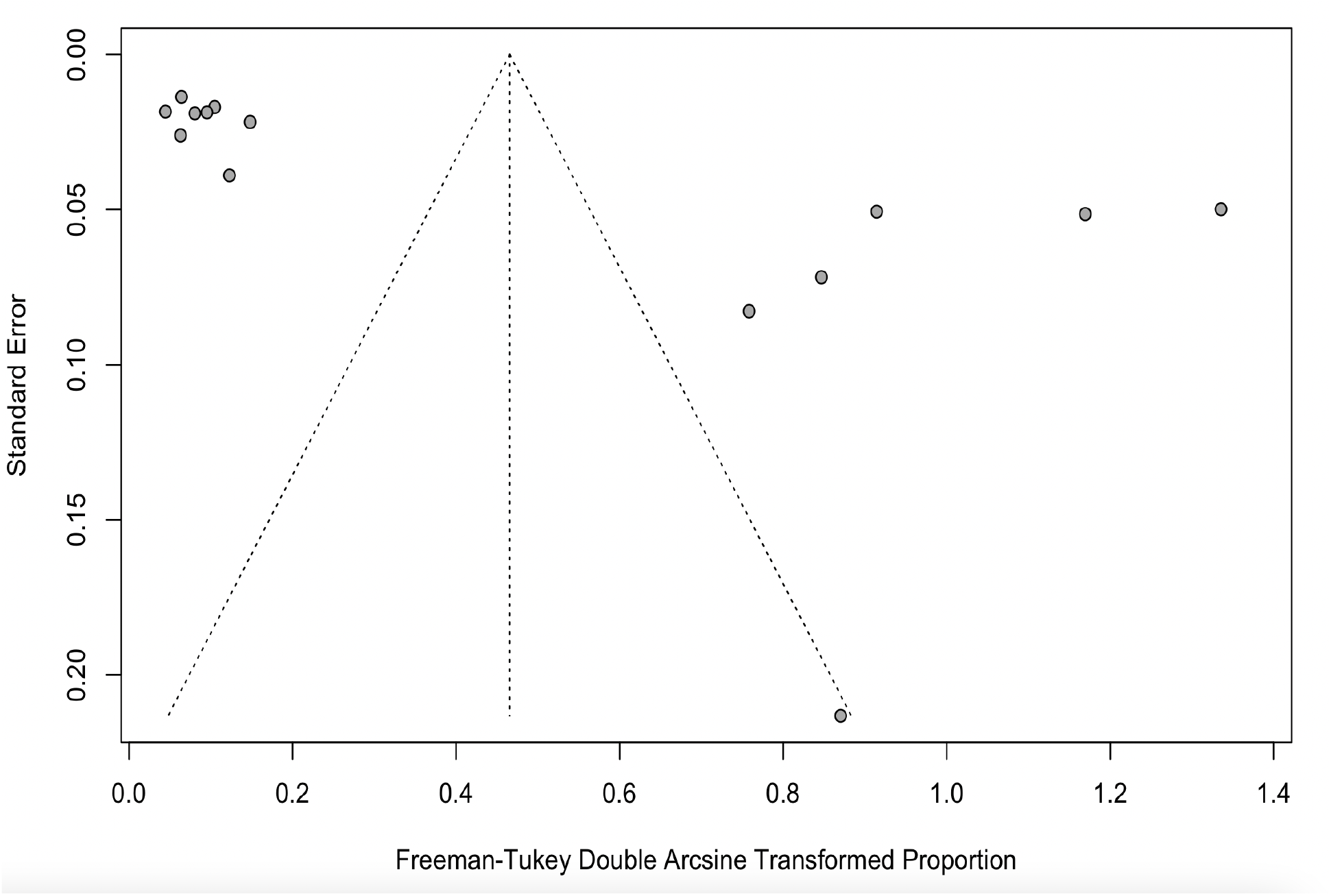
The Funnel plot of CRAB studies shows the publication bias of the study sample

**Fig. 6.**
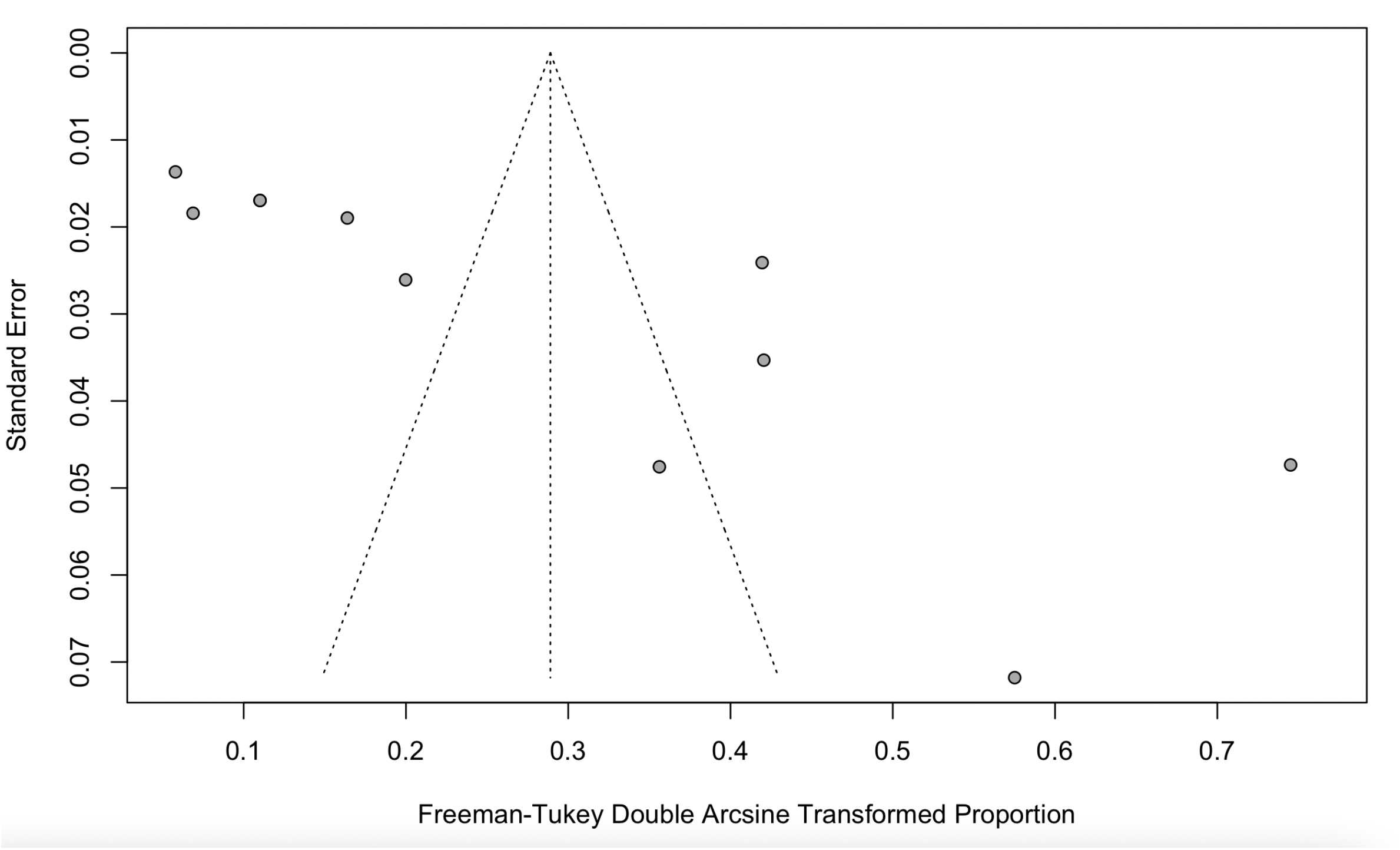
The Funnel plot of CRPA studies shows the publication bias of the study sample

**Figure 6.**
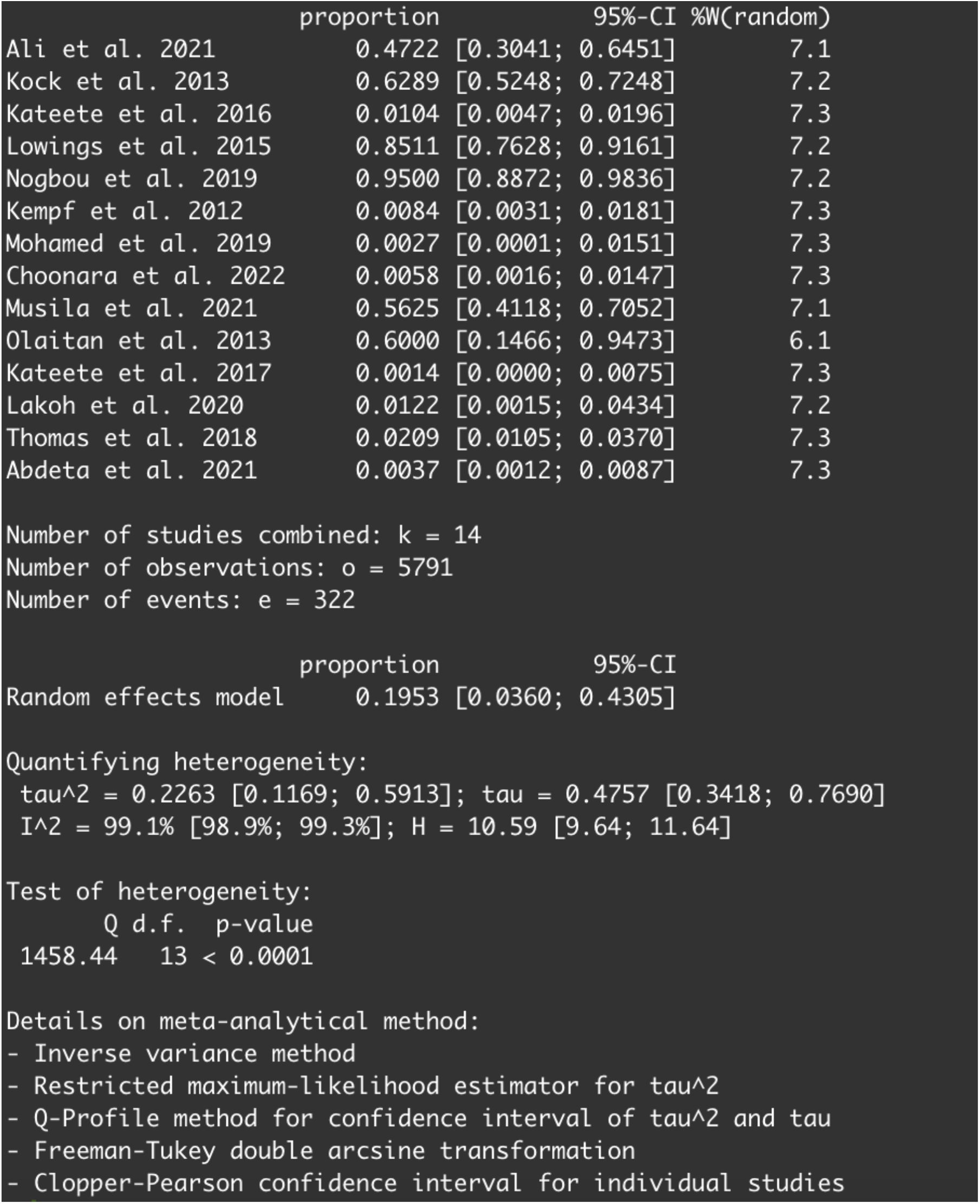
Analysis summary for CRAB

**Figure 7.**
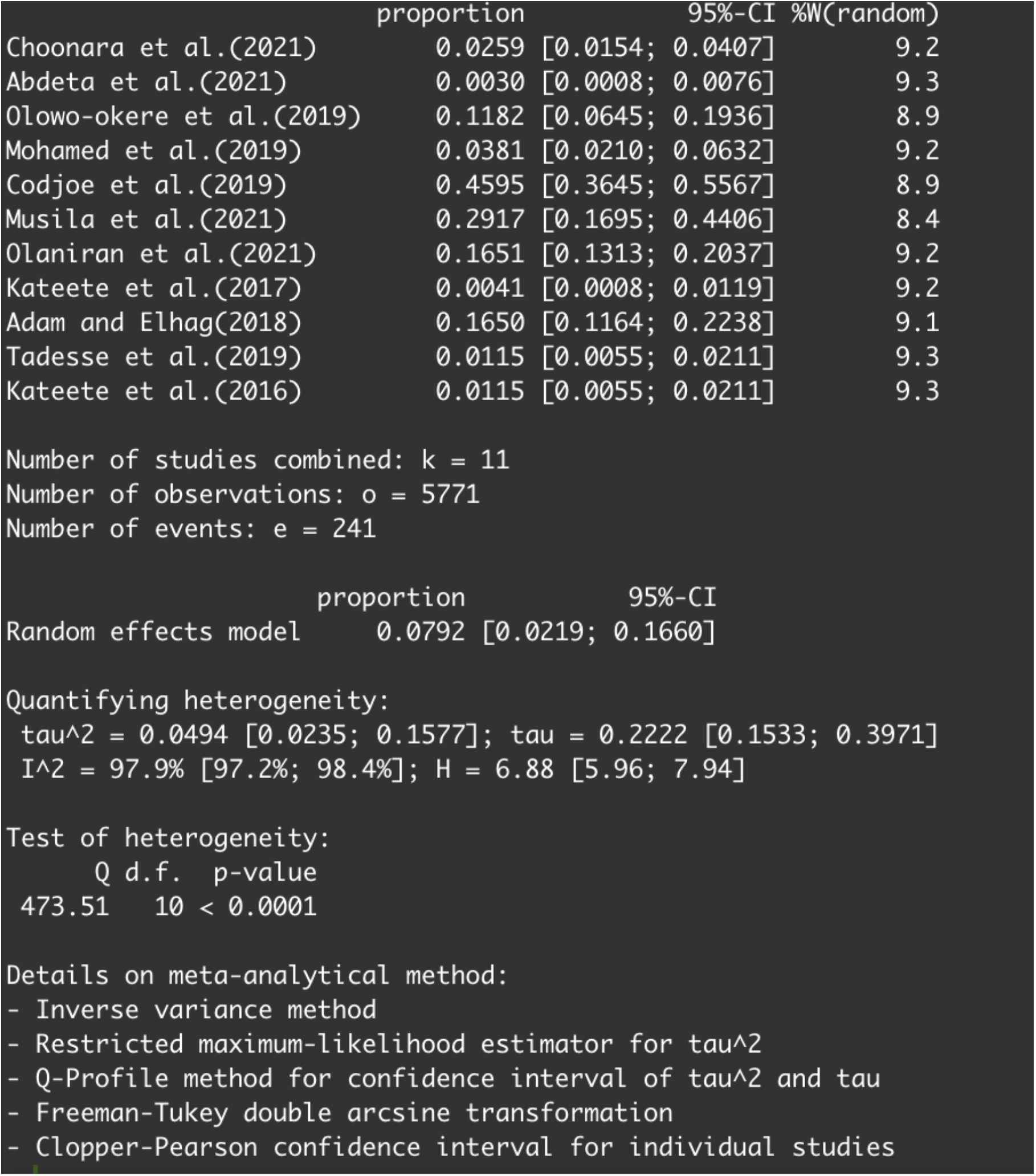
Analysis summary for CRPA

### Prevalence of Carbapenem-resistant genes in *A. baumanii* and *P. aeruginosa* in sub-Saharan Africa

The carbapenemase genes isolated from *A. baumanii* reported in the articles analysed include bla_OXA23,_ bla_OXA48_, bla_GES._, bla_NDM,_ bla_VIM_, bla_OXA24_, bla_OXA58_, bla_OXA51_, bla_SIM-1_, bla_OXA40_, bla_OXA66_, bla_OXA69_, bla_OXA91_. The most common carbapenemase gene in the studies analysed are the bla_OXA23_ and bla_VIM_. On the other hand, the carbapenemase genes reported in P. aeruginosa from studies in sub-Saharan Africa included in this analysis are bla_NDM,_ bla_VIM_, bla_IMP_,, bla_OXA48_, bla_OXA51_, bla_SIM-1_, bla_OXA181_, bla_KPC_, bla_OXA23_, bla_OXA50_. The most frequent among them are the bla_VIM_ and bla_NDM_ genes.

**Table 2.** Data extraction table for carbapenem-resistant *P. aeruginosa* studies.

| First author | Year | Study design | Study country | Study aim | Sample size | Sample source | *P. aeruginosa* prevalence | Carbapenem-resistant *P. aeruginosa* prevalence (%) | AST method | Carbapenem used |
| --- | --- | --- | --- | --- | --- | --- | --- | --- | --- | --- |
| Choonara *et al.* | 2022 | cross sectional | Malawi | To ascertain antimicrobial resistance (AMR) in clinical bacterial pathogens from in-hospital adult patients at a tertiary hospital in Malawi | 694 | human | 29 | 18 (2.5%) | Disc diffusion | Meropenem |
| Abdeta *et al.* | 2021 | cross sectional | Ethiopia | To detect and phenotypically characterise carbapenemase no-susceptible Gram-negative bacilli at the Ethiopian Public health institute | 1337 | human | 36 | 4(0.2%) | Disk diffusion | Meropenem |
| Olowo-okere *et al.* | 2019 | cross sectional | Nigeria | To investigate the occurrence of MBL-producing bacteria in a healthcare facility in Nigeria. | 110 | Human |  | 13(11.8%) | Combined disc method |  |
| Mohamed *et al.* | 2019 | cross sectional | Sudan | To characterise the phenotypes and genotypes associated with carbapenems-resistant in gram-negative bacilli isolated | 367 | human |  | 14(3.8%) | Disk diffusion | Meropenem |
| Codejoe *et al.* | 2019 | cross sectional | Ghana | To apply phenotypic and genotypic methods to identify and characterise carbapenem-resistant (CR) Gram-negative bacteria from the hospital environment in Ghana. | 111 | Human |  | 51(45.9%) | Disc synergy | Edoripenem (75%), imipenem (66.7%) and meropenem (58%) |
| Musila *et al.* | 2021 | Cross-sectional | Kenya | To understand the antibiotic resistance profiles, genes, sequence types, and distribution of carbapenem-resistant Gram negative bacteria from patients in six hospitals across five Kenyan counties by bacterial culture, antibiotic susceptibility testing and whole genome sequence analysis. Culture, antibiotic susceptibility testing, and whole-genome sequence analysis. | 48 | Human | 14 | 14(29.2%) |  | Meropenem |
| Olaniran et al. | 2021 | cross sectional | Nigeria | To determine the incidence and Molecular Characterization of Carbapenemase Genes in Association with Multidrug-Resistant Clinical Isolates of *Pseudomonas aeruginosa* from Tertiary Healthcare Facilities in Southwest Nigeria | 430 | human | 430 | 71(16.5%) | Disc difussion | Meropenem and doripenem |
| Kateete et al. | 2017 | cross sectional | Uganda | To report the intra-species genotypic diversity among *P. aeruginosa* and *A. baumannii* isolated from hospitalized patients and the environment at Mulago Hospital, using repetitive elements-based PCR (Rep-PCR) genotyping. | 736 | Human | 9 | 3(0.4%) |  | Meropenem |
| Adam and Elhag | 2018 | Cross sectional | Sudan | To detect (blaVIM, blaIMP and blaNDM) Metallo-β-lactamase genes in Khartoum state | 200 | Human | 54 | 33(16.5%) | Disc diffusion | Meropenem and/ or imipenem. |
| Tadesse *et al.* | 2019 | Cross sectional | Ethiopia | To identify and determine multi-drug resistant, extended spectrum β-lactamase and carbapenemase producing bacterial isolates among blood culture specimens from pediatric patients under five years of age from Tikur Anbessa Specialized Hospital using an automated BacT/Alert instrument. | 340 | human | | 4(1.1%) | Disk diffusion | Meropenem |

CRAB = Carbapenem-resistant *Acinetobacter baumani*
CRPA = Carbapenem-resistant *Pseudomonas aeruginosa*
\*The percentage in the bracket is a fraction of the sample size

## DISCUSSION

Carbapenems are important broad-spectrum antimicrobials of last-resort, hence, resistance to them signifies increased infection mortality, hospital stay duration, and cost of treatment (Friedman et al., 2016). More importantly, when the infection is associated with notorious clinical pathogens such as *A. baumanii* and *P. aeruginosa*. The two Gram-negative pathogens have been linked to varieties of hospital-acquired infections and multidrug resistance. The true prevalence of these pathogens in sub-Saharan Africa is not well known, especially the carbapenem-resistant strains. This study was carried out to determine the prevalence of the carbapenem-resistant strains of *A. baumanii* and *P. aeruginosa*.

Carbapenemase-resistant *A. baumannii* in Sub-Saharan Africa in this study was estimated at 20% (95% CI; 0.04 – 0.43; I^2^=99%; P <0.01). Olaniyi et al. (2019) in their study on the incidence and prevalence of hospital-acquired (carbapenem-resistant) *A. baumannii* in Europe (EUR), Eastern Mediterranean (EMR) and Africa (AFR) reported similar findings with a pooled incidence of Hospital Acquired-CRAB of 21.4 (95% CI 11.0-41.3) cases per 1,000 patients in the EUR, EMR and AFR WHO regions. On the other hand, our study revealed the pooled prevalence of CRPA in sub-Saharan Africa to be 8% (95% CI; 0.02 – 0.17; I^2^=98%; P <0.01). This value is relatively low compared to the prevalence of carbapenem resistance reported in Indonesia, India, Italy, China, Germany, and Spain by Wang et al. (2021). Another study from Asia reported a prevalence of 18.9% CRPA in the Asia-Pacific region. The surprisingly lower prevalence reported in this study might be due to the absence data from many of the sub-Saharan African countries. The high heterogeneity observed in this study is most likely not due to publication bias but a result of the different prevalence rates reported in the different studies analysed from different parts of sub-Saharan Africa and the low number of eligible articles analysed in this study.

The most common carbapenem-resistant genes in *P. aeruginosa* in the compared studies in this analysis are the NDM and VIM genes. In the reports of Wang et al. (2021) the *bla*_VIM_ and *bla*_IMP_ were the most common carbapenemase genes in *P. aeruginosa* in Italy and Indonesia. The dominance of OXA-23 genes in CRAB isolates in many of the studies is not surprising as this gene has been associated with carbapenem resistance in this organism since the 1980s (Evans and Amyes, 2014). Furthermore, an earlier study showed that the presence of plasmid encoding OXA-23 alone in *A. baumanii* is enough to confer resistance to carbapenem in *A. baumanii* (Heritier et al., 2005). Even though some of the studies analysed reported carbapenemase genes, many of them did not report the molecular basis of carbapenem resistance in their study. This might be a result of low resources for molecular technics in many sub-Saharan African labs.

The pooled prevalence during 2012–2022 should maximally reflect the current status of antibiotic resistance. Thus, we believe that the current prevalence of antibiotic resistance in *A. baumannii* infection is similar to the Sub-Saharan countries where these studies were carried out, most of which are facing the same degree of severity of antibiotic resistance. Most published studies have concentrated on the hospital epidemiology of these organisms and animal health care settings making it difficult to demonstrate the extra-hospital origin of *A. baumannii*. Thus, although antibiotic resistance has long been considered as a modern phenomenon, it predates the concept of selective antibiotic pressure due to clinical antibiotic usage (D’Costa *et al*., 2011).

Over the last few decades, the intensive use of antibiotics in humans and as growth-promoting and as prophylactic agents in livestock have resulted in serious environmental and public health problems since this enhances antimicrobial selective pressure (Anane *et al*., 2019). According to a study by Gros et al. (2006), it was discovered that A. *baumannii* isolated from various environmental locations has been linked to nosocomial spread. The resistant bacteria from the extra-hospital environments may be transmitted to humans, to whom they cause diseases that are difficult to treat with conventional antibiotics.

A major strength of our study is that we included studies comprising of non-disease-specific patients from patients, which ensured the representativeness of our estimates for these institutions. However, our study has some limitations. Firstly, the country representativeness of the individual studies is unclear in most cases, limiting our findings’ external validity. Secondly, studies are not evenly distributed across the Sub-Saharan regions. Some countries have more studies included in the analysis while there were no studies from others. Consequently, possible differences in CRAB and CRPA incidence and prevalence between the countries may be masked by geographical proximity. Thirdly, due to the relatively low number of hospital-wide studies, our hospital-wide estimates of hospital-acquired *A. baumannii* infections are unlikely to be generalizable.

## CONCLUSION

*Acinetobacter baumannii* and *P. aeruginosa* constitute two important pathogens of public health concern due to their increasing multidrug-resistant nature. The prevalence of CRAB and CRPA reported in sub-Saharan Africa indicates a serious threat to the sub-Saharan African populace. Sensitization on the dangers of self-medication, and poor antibiotics stewardship should be intensified as well as advocacy for the reduction in the use of antibiotics for growth promotion and prophylactics in animals. Urgent epidemiological studies through comprehensive surveillance of the pathogen at both hospital and community-based that will showcase the prevalence of CRAB in the environment, animal/animal products, and the hospital is advocated to be conducted with the inclusion of sub-Saharan African countries currently lacking this data. Moreover, Finally, infection control policies promoting personal and environmental hygiene, and appropriate administration of antibiotics by clinicians and veterinarians should be made and enforced to facilitate the reduction of CRAB in the region.

## Supporting information

Search strategy and eligibility criteria

## Data Availability

All data produced in the present work are contained in the manuscript

