## Supplementary material for "Prevalence Of carbapenem resistance in *Acinetobacter baumanii* and *Pseudomonas aeruginosa* in sub-Saharan Africa: a systematic review and meta-analysis": Search strategy and eligibility criteria

Search strategies

Five major databases (Google scholar, PubMed, Scopus, Africa Journal Online, Web of Science) were searched for articles used in this study. The keywords and search terms used include; Carbapenem AND resistant OR resistance AND Acinetobacter baumanii OR A. baumanii/ AND/OR Pseudomonas aeruginosa/ OR P. aeruginosa AND sub-Saharan Africa OR SSA OR country names. The last search was on the 31st of July 2022.

**Eligibility criteria**

Inclusion criteria

- Articles published between 2012 and 2022
- Cross-sectional studies
- Articles that reported carbapenem-resistance in *P. aeruginosa* and/or *A. baumanii*
- Only studies that reported isolates from human

Exclusion criteria

- Literature Reviews, systematic reviews, and meta-analysis
- Articles published before 2012
- Articles reporting prevalence outside sub-Saharan Africa
- Articles that did not screen for carbapenem resistance in these organisms.
- Articles that did not specify the species of interest
- Articles with no reported case of carbapenem resistance in these organisms
